# Multiple Cost Optimisation for Alzheimer’s Disease Diagnosis

**DOI:** 10.1101/2022.04.10.22273666

**Authors:** Niamh McCombe, Xuemei Ding, Girijesh Prasad, David P. Finn, Stephen Todd, Paula L. McClean, KongFatt Wong-Lin, the Alzheimer’s Disease Neuroimaging Initiative

## Abstract

Current machine learning techniques for dementia diagnosis often do not take into account real-world practical constraints, which may include, for example, the cost of diagnostic assessment time and financial budgets. In this work, we built on previous cost-sensitive feature selection approaches by generalising to multiple cost types, while taking into consideration that stakeholders attempting to optimise the dementia care pathway might face multiple non-fungible budget constraints. Our new optimisation algorithm involved the searching of cost-weighting hyperparameters while constrained by total budgets. We then provided a proof of concept using both assessment time cost and financial budget cost. We showed that budget constraints could control the feature selection process in an intuitive and practical manner, while adjusting the hyperparameter increased the range of solutions selected by feature selection. We further showed that our budget-constrained cost optimisation framework could be implemented in a user-friendly graphical user interface sandbox tool to encourage non-technical users and stakeholders to adopt and to further explore and audit the model - a humans-in-the-loop approach. Overall, we suggest that setting budget constraints initially and then fine tuning the cost-weighting hyperparameters can be an effective way to perform feature selection where multiple cost constraints exist, which will in turn lead to more realistic optimising and redesigning of dementia diagnostic assessments.

**Clinical Relevance:** By optimising diagnostic accuracy against various costs (e.g. assessment administration time and financial budget), predictive yet practical dementia diagnostic assessments can be redesigned to suit clinical use.

## I. Introduction

Dementia is exacerbated by gradually ageing societies and the current sub-optimal dementia care pathway [1]. The latter impacts everything from diagnosis to management of care (e.g. [2], [3]). Cognitive and functional assessments (CFAs) form a key component of the dementia diagnosis process within the clinical care pathway [4]. However, CFAs can vary in terms of diagnostic accuracy, sensitivity and specificity [4], and they are not always administered in a standardised way across clinical practices [1]. In addition to the difficulty of diagnosing dementia, clinicians’ consultation time is also typically limited, within the order of minutes [5], [6], [7]. This opens up opportunities for machine automation to aid dementia diagnosis [1].

Recently, there has been a rapid increase in research activities in dementia data science, especially on applying machine learning to classify (‘diagnose’) the severity of dementia, particularly Alzheimer’s disease (AD), which is the most common dementia type [1], [5] This has been facilitated by openly available dementia datasets [1]. Such open datasets, together with longitudinal and clinical datasets, often come with large number of variables (i.e. data features), and hence feature selection is often used [8].

A practical use of feature selection would be to guide, as clinical decision support, effective clinical diagnosis using only a small subset of data features [9]. However, although feature selection has been used extensively in dementia data science [6], [9], [10], [11], the practical costs of dementia assessments have not been put into much consideration. As far as we know, within dementia data science, only one previous study has been identified that incorporated cost-sensitive feature selection methods, penalising CFA features that take longer to administer in the evaluation criteria [7]. A limitation of this work was that the hyperparameter used to weight the cost function in the feature selection method is non-intuitive and has no clear and obvious meaning to the user. Furthermore, the data used in that study were CFAs and demographics, but in clinical settings additional biological features may also be involved as part of the diagnostic assessment [1]. More importantly, additional types of costs (e.g. financial budgets) were not considered. For instance, the most accurate diagnostic methods may not be cost-effective if they involve, for example, very expensive neuroimaging scans. Hence, in more realistic budget-constrained situations, budget threshold(s) should be incorporated into the feature selection optimisation process.

In this work, we formally generalise a previous cost-sensitive approach [12] to multiple cost types, while setting maximal total costs (e.g. maximal total cost budget). We then apply this generalised algorithm to an open dementia dataset, incorporating not only CFAs, but also various types of neuroimaging and other biomarker data. We then show how the algorithms operate in practice by using both assessment time and financial budget as examples for the different cost types. The effects of cost hyperparameters and maximal costs on the classification model performance will be investigated. Further, to encourage non-technical stakeholders to be more involved in the adoption of our proposed model, our algorithms are embedded within a user-friendly graphical user interface (GUI) such that specific cost values and constraints, and thus, the model can be edited or updated.

## II. METHODS

### A. Data Description and Cost Estimation

We used the dataset generated by [7] for our feature selection. The authors used the Alzheimer’s Disease Neuroimaging Initiative (ADNI) open datasets (adni.loni.usc.edu) and combined CFA sub-features (specific queries) from Mini Mental State Examination (MMSE) [3], Montreal Cognitive Assessment (MoCA) [13], Alzheimer’s Disease Assessment Scale (ADAS) [2], Functional Activities Questionnaire (FAQ) [14], Everyday Cognition – Patient scale [15], Geriatric Depression Scale [16], and Neuropsychological Battery (NB) and used multiple methods of feature selection to generate a set of 35 sub-features which were frequently selected, and assigned time costs in seconds to these sub-features [7]. In this dataset, Clinical Dementia Rating Sum-of-Boxes (CDR-SB) rating was used as a measure of dementia severity [7]. Following [7], CDR-SB was also re-coded into 3 categories: Cognitively Normal (CN); Mild Cognitive Impairment (MCI); and Alzheimer’s Disease (AD) (comprising mild, moderate and severe AD cases).

In our study, high-level neuroimaging and cerebrospinal fluid based biomarker data was added to the dataset as proof of concept to investigate feature selection under multiple budget constraints. Financial costs for acquiring these biomarkers were estimated from the literature [17], [18], [19]. Detailed information about the data features, including biomarker features, and their associated time and financial costs, is shown in Table I in the Appendix. Note that we did not consider waiting time, technician time, data acquisition time, and biomarker data analytical time when considering the biomarkers. Besides, such timescales are usually much longer than the assessment times for administering the CFAs; these time costs are not fungible with the time costs for administering CFAs, which must take place within a brief clinician appointment. Hence, for simplicity and clarity, we set the assessment time costs of all biomarker assessment items to be zero. In a similar vein, the financial costs for CFAs will generally be much smaller than the biomarkers. Hence, we set the financial costs of all CFA assessment items to be zero.

### B. Cost-sensitive Feature Selection Methods

#### Cost-sensitive correlation-based feature selection (CFS)

Correlation-based feature selection (CFS) [20], [21] evaluates the worth of every possible subset of features by considering the individual predictive ability of each feature along with the degree of redundancy between them, with a best-first-search algorithm through the feature space (see Algorithm 1 below). The CFS algorithm finds an optimal set of features which correlate with the class variable and do not correlate with each other. It uses a “Merit” heuristic to evaluate the best feature set, described by

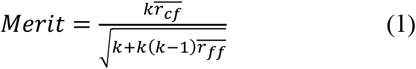

where *k* is the number of features in the set, 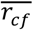 is the average feature-class correlation, and 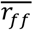 is the average feature-feature correlation. In the implementation in the FSelector package [21], the correlation between discrete features is measured by mutual information [22], and for continuous features the correlation coefficient is used. It can be seen that the merit heuristic incorporates a penalty to favour smaller sets of features over larger sets. The function then uses a best-first search to find the set of features with the highest merit. See Algorithm 1 below.

##### Algorithm 1

CFS Using Best-First-Search Algorithm

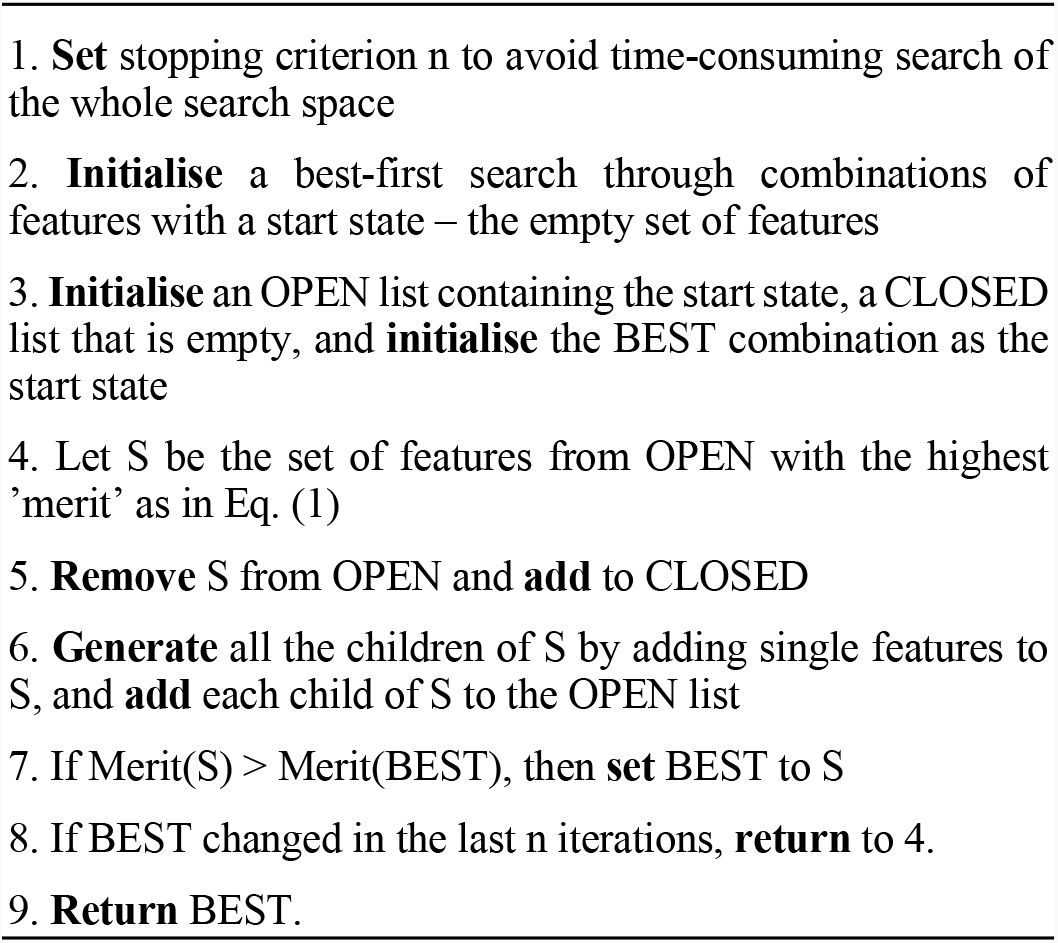

The cost-sensitive feature selection framework laid out in [12] was used to modify the CFS implementation in Fselector [21] to incorporate feature costs as detailed in [7]. Cost-sensitive CFS incorporates a cost weighting parameter, *λ*, which can be varied to reflect different values of cost weighting for the features. If *λ* is set to zero, the algorithm performs like a typical CFS without (time) cost. This modification for cost sensitivity can be generalised to any feature selection algorithm which has an evaluation function. We used CFS here as it performed better than other feature selection algorithms in other work with the same dataset [7].

Cost-sensitive CFS [12] extends the CFS algorithm by adding a cost penalty to the Merit function as shown:

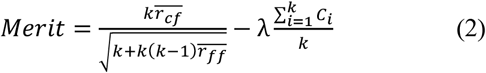

where *C*_*i*_ is the feature cost of item *i*, which in this case is the time of each assessment item, and λ is the cost weighting parameter which can be varied to increase or decrease the importance of feature cost in the merit heuristic.

#### Cost-sensitive CFS with budget threshold

In this work, we have adapted cost-sensitive CFS further to incorporate a budget constraint as shown:

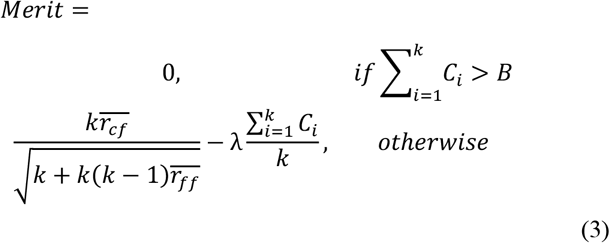

where *B* is the budget threshold for some cost type. This means that feature sets which exceed the budget threshold are simply not evaluated. If there are instead *N* (*>* 1) types of cost, then the merit function can be generalised as follows:

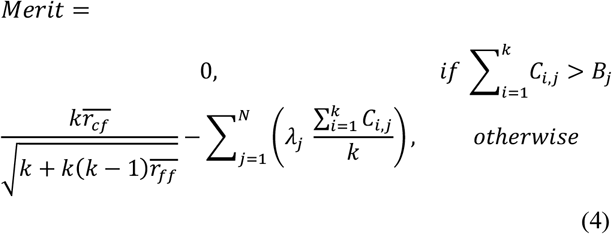

for any cost type *j*.

### C. Data Modelling and Testing

The algorithm in Section II.B was implemented in R by adapting the code from the FSelector package [21]. The parameter n in the best-first search was increased from the default of 5 to 14 for more thorough exploration of the search space. The maximum and minimum values of the cost weighting hyperparameters *λ*_1_ and *λ*_2_, corresponding to the two cost types in the data, were ascertained through trial and error. The budget thresholds were disabled for initial exploration of the data, and *λ*_1_ and *λ*_2_ were varied from maximum to minimum in a stepwise manner, for a total of 176 iterations of the algorithm with different *λ*_*j*_ values.

For validating the selected item sets, the data was split into 80% training data and 20% testing data. A random forest model for each selected feature set was built and tuned using 10-fold cross-validation on the training data. The resultant models were then tested on the withheld testing data. The multiclass receiver operating characteristics (ROC) area-under-curve (AUC) of these models was then calculated by the process defined in [23] using the pROC package [24]. This method of calculating multiclass AUC extends the binary AUC concept to multiple classes (in our case, 3 classes) by calculating the pairwise AUCs of each class against every other class and then averaging the results.

Both budget constraints were varied in tandem. *B*_1_ was assigned values from 100 to 2000 in intervals of 100 in turn. *B*_2_ was assigned the values (0, 250, 500, 750, 1000, 2000, 2250, 2500, 2750, 3000) – every possible combination of financial costs in this dataset. Thus, the algorithm was run 200 times for each selected value of *λ*_1_. At a later part of the evaluation, the *λ*_2_ parameter for financial costs was set 135 to 0 throughout (due to its erratic relationship with financial budget). The *λ*_1_ parameter for assessment time costs was set to 0 for the first iteration of this experiment, and then the experiment was repeated twice more, once with *λ*_1_ set to 0.004 (the value associated with the median selected item set) and once with *λ*_1_ set to 0.009 (the largest value that produced an item set with an associated AUC above 0.85). The model AUC associated with each item set selected in this process was calculated as above.

### D. GUI Development

The method for estimating assessment item time costs used in [7] can be refined for future use through humans-in-the-loop approach. For instance, clinicians who work with dementia patients may have access to their own estimates of assessment times. It may also be the case that the financial costs of different dementia assessments may vary between different locations [25]. We developed a GUI-based sandbox-like toolbox using the abovementioned optimisation algorithms to allow non-technical users to input their own cost estimations and budget thresholds, build a new model, and obtain a new optimal set of diagnostic assessments that suit their situation and use. We implemented the GUI in R Shiny [26] using the DT package in R [27] to create an editable data table.

## III. RESULTS

As proof of concept for our proposed generalized algorithm as described in Section II.B, we vary the cost weighting parameters, *λ*_1_ and *λ*_2_, and the budget thresholds, *B*_1_ and *B*_2_, where subscripts 1 and 2 denote assessment time cost and financial cost, respectively. This will help determine whether the multiple-cost budget-constrained version of the cost-sensitive CFS algorithm is effective and practical.

### A. Relationship between Assessment Time, Financial Cost and Classifying AD Severity

With no budget threshold set, most of the selected item sets were highly predictive (high AUC values) of classifying AD severity (CN, MCI or AD), as can be seen in Fig. 1. Moreover, as financial cost decreased (lighter colours), assessment time increased while AD classification AUC increased before saturating at around 0.9. Hence, there was a trade-off between financial cost and assessment time, and between assessment time and classification AUC. The most accurate item set, consisting only of CFA values, achieved a multiclass AUC score of 0.9 (Fig. 1, farthest top-right corner), although this item set was very costly in terms of assessment time duration. Further, although the algorithm was run 176 times with different values of the *λ*_*j=*1,2_ hyperparameters, only 18 unique item sets were selected. It turned out that most of the item sets incorporated both CFA values and biomarker features. However, 3 item sets incorporated no CFA values, and had limited predictive value (Fig. 1, ∼0 assessment time), and 3 item sets incorporated only CFA values (Fig. 1, light yellow squares, with assessment times of 538, 568 and 1764 seconds).

**Figure 1.**
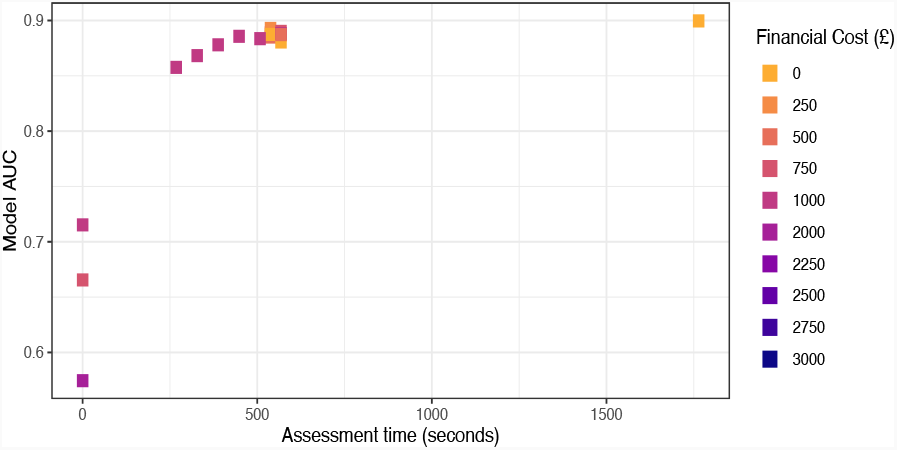
AUC of selected item sets by assessment time cost and financial cost (legend).

### B. Budget Constraints More Intuitive than Cost Weighting Parameters

After investigating the interplay between AD-classification AUC, assessment time and financial cost, we next seek to understand how the hyperparameters *λ*_*j=*1,2_ can affect the total assessment time and total financial cost. We found that varying the *λ*_1_ hyperparameter for the assessment time cost budget generally led to a decrease in the total assessment time for selected CFA items, but the relationship was nonlinear nor well predictable (Fig. 2a). Similarly, by varying the *λ*_2_ hyperparameter for the financial cost budget, we found no apparent trend with regards to the total financial costs in the selected item set (Fig. 2b). Thus, while the use and value of cost-sensitive feature selection was apparent from the results, the *λ* hyperparameters for cost weighting were imprecise and inefficient to optimise the algorithm, especially when multiple budget constraints must be considered.

**Figure 2.**
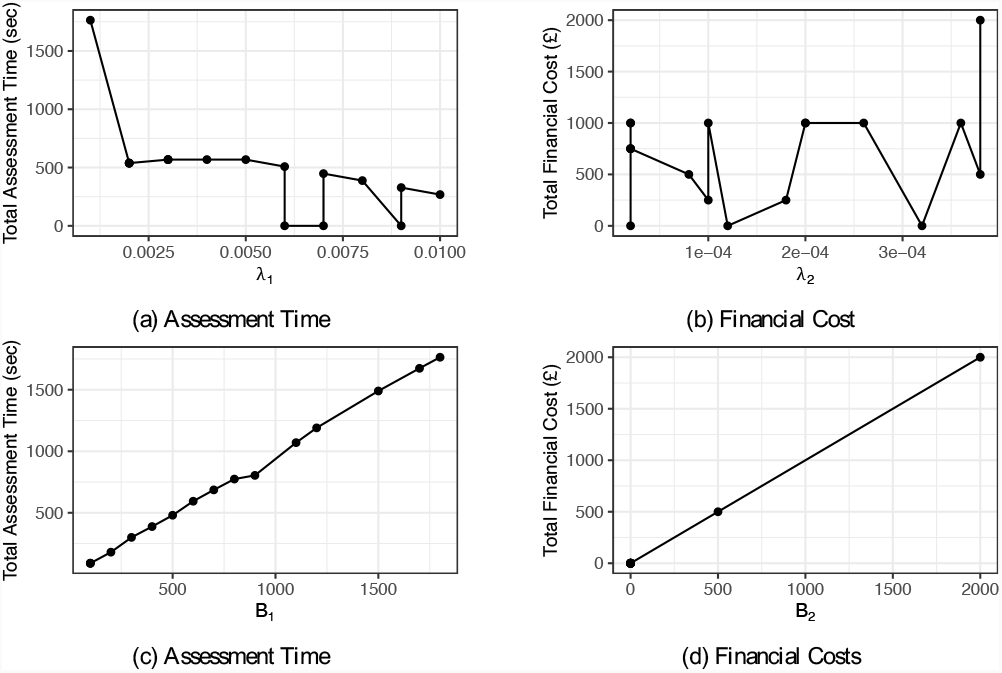
Evaluation of total assessment time and total financial cost with respect to their corresponding hyperparameters *λ*_1_ and *λ*_2_, and *B*_1_ and *B*_2_. (a) Total assessment time cost generally decreases when the cost-weighting hyperparameter *λ*_1_ increases, but the relationship is not monotonic but fluctuates. (b) Almost random relationship betwen total financial cost and cost-weighting hyperparameter *λ*_2_. (c) There is an almost one-to-one linear relationship between between the *B*_1_ value supplied by the user and the total assessment time cost. (d) The relationship between *B*_2_ and the total financial cost is exact and direct.

Instead of using the hyperparameters *λ*_*j*_ to control the feature selection algorithm, we now used the total budget thresholds, varying *B*_1_ from a minimum of 100 to a maximum of 2000, and varying *B*_2_ from a minimum of 0 to a maximum of 3000. We found that it was possible to control the algorithm easily and intuitively. In particular, the algorithm consistently returned results close to the user-specified budget threshold on both budgets (Figs. 2c and d). In fact, there is an almost one-to-one linear relationship between user-specified total time (financial) budget and the selected total time (total financial cost).

### C. Cost Weighting Parameters Provide Variability in Assessment Time, Classification Accuracy and Feature Types

As the *λ*_2_ hyperparameter for adjusting sensitivity to financial costs appeared to have little effect on the algorithm, as shown in Fig. 2b, this parameter was set to 0 and not investigated henceforth. To examine the impact of the budget threshold parameter, the budget thresholds for both cost types were varied simultaneously in a stepwise manner as described in Section II.C. We then varied hyperparameter *λ*_1_ with respect to the assessment time costs and model classification.

As illustrated in Fig. 3, we found *λ*_1_ to have a strong effect on both the classification AUC and assessment time. For instance, when *λ*_1_ was set to 0 (Fig. 3, circles), the algorithm found relatively few item sets that fit within the time budget constraints (18 unique item sets were found from 200 variations of budget constraints, all of which consisted only of CFA items). At the other end, when *λ*_1_ was set to a high value of 0.007 (Fig. 3, squares) again relatively few item sets were found (9 item sets) and with the exception of one instance, which selected a single CFA assessment (RAV LT:immediate), only biomarker features were selected.

**Figure 3.**
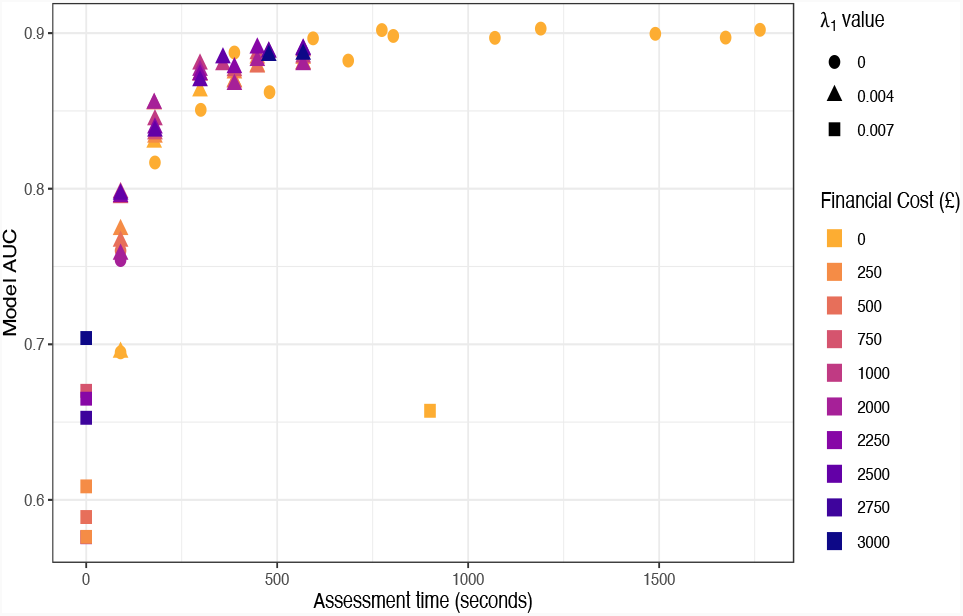
AUCs and associated costs of budget-constrained feature sets, with values of *λ*_1_ set to 0 (circles), 0.004 (triangles,) and 0.007 (squares), and *λ*_2_ set to 0. The relationship between budget constraints specified by user and the associated costs is shown in Fig. 2.

With an intermediate value of *λ*_1_(= 0.004), the algorithm generated a larger selection of item sets, selecting 50 different item sets containing both neuromarker and CFA assessments, out of the 200 variations of budget constraints that were tested. Therefore, even when budget constraints are specified, there is an important role for the cost-weighting hyperparameters. Non-optimal values of these parameters produce limited options for the user, as shown by the circle and square markers in Fig. 3. The intermediate value of *λ*_1_ appears to introduce more variety into the feature selection procedure. It should also be noted that without budget constraints, when only *λ*_1_ and *λ*_2_ were varied, 18 distinct feature sets were selected from 176 iterations of the algorithm. Here, 50 distinct sets were selected from 200 iterations of the algorithm. The introduction of budget constraints has also increased the variety of selected item sets.

It should be noted that the range of AUCs of feature sets selected under the budget-constrained condition were comparable to the range of AUCs shown in Fig. 1. Hence, it is reasonable to conclude that the presence of the budget condition imposes little penalty on the effectiveness of the feature selection algorithm.

Unsurprisingly, some of the selected feature sets produced much greater diagnostic accuracy, as measured by multiclass AUC, than others. The four biomarker and neuromarker features, when not combined with CFA items, were not diagnostically accurate (AUC of 0.72) with an associated financial cost of 3000 (Fig. 3, top left square) The maximal AUC of 0.902 was achieved when *λ*_1_ was set to 0, i.e. no cost weighting on assessment time. Here, an item set consisting only of CFA features with an associated total assessment time of 1764 seconds (24 minutes) was selected (Fig. 3, top right circle.) When assessment time was considered by the feature selection evaluation function, with *λ*_1_ set to 0.004, (Fig. 3, triangles) the maximal AUC achieved was 0.890, with an associated assessment time cost of 448 seconds and financial cost of 2250 (top-most purple triangles with assessment time of 448 seconds).

### D. User-friendly GUI for Humans-in-the-loop Modelling

Based on the GUI application and associated code developed by [7], and licensed under the Academic Free License 3.0, we developed a user-interface GUI for selecting diagnostic items under multiple budget constraints. This software application is made available at https://niamh-m.shinyapps.io/DementiaPathway/.

An overall view of the application is shown in Figure 4a. The application is navigated through a tabbing system. Upon opening the application, there is a tab that displays a selection box (Fig. 4a, right) which allows users to choose any subset of the assessment items in our dataset for analysis. The cost weighting parameters for assessment time costs and financial costs can be varied with slider inputs (Fig. 4a, left). Alternatively, assessment time constraints and budget constraints may be entered via text inputs within the textboxes (Fig. 4a, left).

**Figure 4.**
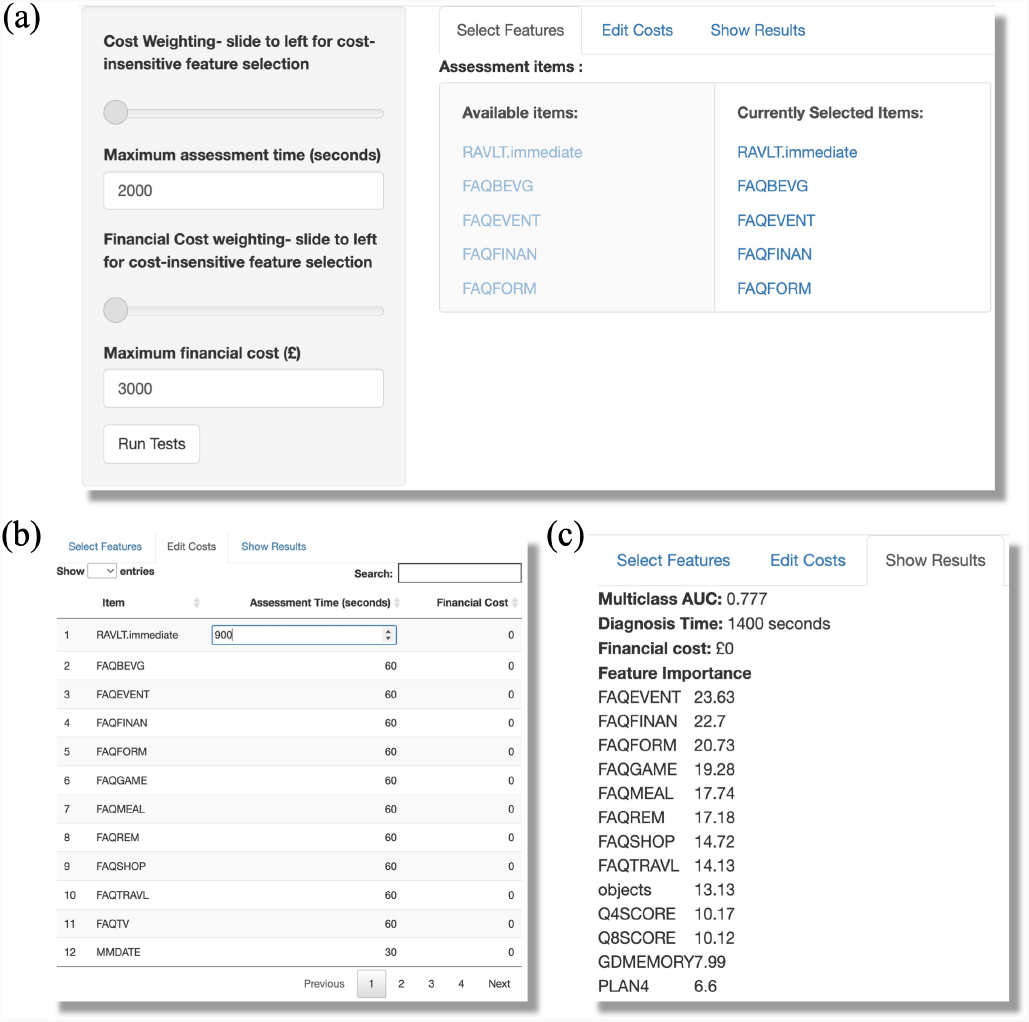
A GUI for stakeholders to perform budget-constrained feature selection. (a)-(c): Steps for editing, selecting and output. (a) Selection of assessment items and adjusting hyperparameters. (b) Edit various costs based on user’s perspective. (c) Display selected features and total costs.

The selected items are displayed in an editable data table (Fig. 4b) in another tab, in which the user can make further edits to the time costs or financial costs associated with these features in accordance with their expertise. Assessment time constraints and budget constraints can be entered via text inputs within a textbox. The algorithm is activated with a “Run Tests” button (Fig. 4a, left). Feature selection is performed using user-chosen set of parameters. The resulting set of features are used to build an RF classifier on a random 80% of the training data. The multiclass AUC of the model will be calculated on the remaining 20%. Finally, the AUC, total assessment costs, total financial costs and the feature importance in the RF model of all selected features can then be displayed in the Results tab (Fig. 4c).

## IV. DISCUSSION

On the one hand, many machine learning algorithms, especially in dementia data science research, ignore real-world practical constraints [7]. On the other hand, cost-benefit studies in healthcare, particularly on dementia, do not involve machine learning algorithms [25], [28]. In this work, we bridge this gap by developing machine-learning for classifying AD severity incorporating practical, real-world costs and budget constraints.

In this paper, we have generalised an optimisation algorithm for feature selection involving multiple cost types and constraints and demonstrated its effectiveness in classifying AD severity using an open-source data. As proof of concept, we included data from CFAs with associated assessment time costs, and data from neuroimaging and CSF-based biomarkers with associated financial costs. Building on top of this multi-cost-based optimisation algorithm, we have developed a user-friendly GUI sandbox-like tool for non-technical stakeholders to set multiple budget limits and assessment time costs depending on their expertise and financial budgets. As outcome of this tool, the optimal subsets of AD assessment features will be suggested.

Algorithm wise, we have extended a previous framework for cost-sensitive feature selection by [12] and the time-cost assessment approach by [7] and developed a more practical and intuitive algorithm through our proposed budget-threshold implementation. In fact, we propose here that the extended algorithm can most effectively be deployed by specifying the cost budget constraints first and then fine tuning the cost-weighting (*λ*_*j*_) hyperparameters.

It should be noted that in this work, although the maximal AD severity classification (AUC) accuracy was only very slightly higher than that of [7], even with additional data features (neuroimaging markers and CSF-based biomarkers) incorporated, our approach allows the possibility of selecting different data types. For instance, when weight of assessment time costs is very high (e.g. too many patients for consultation) then biomarker features are suggested.

A limitation of this work is the use of coarse-grained neuroimaging. While CFA data outperformed neuroimaging data in this dataset, this result should not be generalised. The CFA data was previously selected using feature selection [7], while instead of using complex neuroimaging data we have used only a single representative value for the MRI and FDG-PET scans, as proof of concept for the multiple cost constraints. It should be noted that diagnosis based on neuroimaging data can be highly accurate [29], [30]. Future work will utilise more detailed neuroimaging data, and a larger selection of CFA assessments. Another key limitation of this work is that group-based feature selection is not used. In more realistic situations, many data features (e.g. all the information associated with a particular neuroimaging scan) can be acquired with a single cost expenditure. To allow for proper cost-sensitive feature selection on neuroimaging and biomarker data, group-based cost-sensitive feature selection will be implemented. Further, data features and their costs may not be independent of each other, as we have assumed in this work, but may have more complex interrelationships.

Finally, we have implemented a GUI sandbox-like tool on top of our proposed algorithm, for easy accessibility by a wide variety of users. In future work, this application tool will be made available to various stakeholders and feedback will be used to further improve the application. As the GUI tool is sufficiently flexible, future work may incorporate other machine learning models including interpretable/explainable algorithms and data types, and extended to other domains (e.g. other diseases). In general, we believe that such tool can provide a platform for bridging the gap between machine learning research and clinical uptake through the humans-in-the-loop route.

## Data Availability

Software application is made available at https://niamh-m.shinyapps.io/DementiaPathway/

## Appendix

**APPENDIX TABLE I.** Features and Associated Costs.

| Feature Name | Description | Time Cost (sec) | Financial Cost (£) |
| --- | --- | --- | --- |
| *Biomarker Features* |  |  |  |
| TAU.bl | Tau levels (CSF) | 0 | 250 |
| ABETA.bl | Amyloid-Beta levels (CSF) | 0 | 250 |
| FDG.bl | Average FDG-PET (angular, temporal, posterior cingulate) | 0 | 2000 |
| Entorhinal.bl | Entorhinal Atrophy (MRI) | 0 | 500 |
| RAVLT.immediate | Rey Auditory Verbal Learning Test - Immediate Total | 900 | 0 |
| *CFA Features* |  |  |  |
| FAQBEVG | Heating water, making a cup of coffee, turning off the stove. | 60 | 0 |
| FAQEVENT | Keeping track of current events. | 60 | 0 |
| FAQFINAN | Writing checks, paying bills, or balancing checkbook. | 60 | 0 |
| FAQFORM | Assembling tax records, business affairs, or other papers. | 60 | 0 |
| FAQGAME | Playing a game of skill such as bridge or chess, working on a hobby. | 60 | 0 |
| FAQMEAL | Preparing a balanced meal. | 60 | 0 |
| FAQREM | Remembering appointments, family occasions, holidays, medications. | 60 | 0 |
| FAQSHOP | Shopping alone (clothes/household necessities/groceries). | 60 | 0 |
| FAQTRAVL | Traveling out of the neighborhood, driving, or arranging to take public transportation. | 60 | 0 |
| FAQTV | Paying attention to/ understanding TV program/book/magazine. | 60 | 0 |
| MMDATE | What is today's date? | 30 | 0 |
| MMMONTH | What is the month? | 30 | 0 |
| MMObjects | Naming Objects - Immediate/Delayed Total (composite variable) | 300 | 0 |
| Q4SCORE | Delayed Word Recall | 266 | 0 |
| Q1SCORE | Word Recall Task | 266 | 0 |
| Q7SCORE | Orientation (date/place etc) | 266 | 0 |
| Q8SCORE | Word Recognition | 266 | 0 |
| Q13SCORE | Number Cancellation | 266 | 0 |
| CATANIMS C | Category Fluency (Animals) - Total Correct | 150 | 0 |
| TRABSCOR | Trails B | 200 | 0 |
| BNTTOTAL | Boston Naming Test- Total Score | 600 | 0 |
| GDMEMORY | Do you feel you have more problems with memory than most? | 28 | 0 |
| LANG6 | Understanding the point of what other people are trying to say. | 60 | 0 |
| LANG9 | Understanding spoken directions or instructions. | 60 | 0 |
| MEMORY1 | Remembering a few shopping items without a list. | 60 | 0 |
| MEMORY2 | Remembering things that happened recently. | 60 | 0 |
| MEMORY3 | Recalling conversations a few days later. | 60 | 0 |
| MEMORY5 | Repeating stories or questions. | 60 | 0 |
| MEMORY6 | Remembering the current date or day of the week. | 60 | 0 |
| ORGAN2 | Balancing the checkbook without error. | 60 | 0 |
| ORGAN3 | Keeping financial records organized. | 60 | 0 |
| PLAN1 | Planning a sequence of stops on a shopping trip. | 60 | 0 |
| PLAN4 | Thinking things through before acting. | 60 | 0 |
| PLAN5 | Thinking ahead. | 60 | 0 |

## Acknowledgments

Data collection and sharing for this project was funded by the Alzheimer’s Disease Neuroimaging Initiative (ADNI) (National Institutes of Health Grant U01 AG024904) and DOD ADNI (Department of Defense award number W81XWH-12-2-0012). ADNI is funded by the National Institute on Aging, the National Institute of Biomedical Imaging, and through generous contributions from the following: AbbVie, Alzheimer’s Association; Alzheimer’s Drug Discovery Foundation; Araclon Biotech; BioClinica, Inc.; Biogen; Bristol-Myers Squibb Company; CereSpir, Inc.; Cogstate; Eisai Inc.; Elan Pharmaceuticals, Inc.; Eli Lilly and Company; EuroImmun; F. Hoffmann-La Roche Ltd and its affiliated company Genentech, Inc.; Fujirebio; GE Healthcare; IXICO Ltd.; Janssen Alzheimer Immunotherapy Research Development, LLC.; Johnson Johnson Pharmaceutical Research Development LLC.; Lumosity; Lundbeck; Merck Co., Inc.; Meso Scale Diagnostics, LLC.; NeuroRx Research; Neurotrack Technologies; Novartis Pharmaceuticals Corporation; Pfizer Inc.; Piramal Imaging; Servier; Takeda Pharmaceutical Company; and Transition Therapeutics. The Canadian Institutes of Health Research is providing funds to support ADNI clinical sites in Canada. Private sector contributions are facilitated by the Foundation for the National Institutes of Health (www.fnih.org). The grantee organization is the Northern California Institute for Research and Education, and the study is coordinated by the Alzheimer’s Therapeutic Research Institute at the University of Southern California. ADNI data are disseminated by the Laboratory for Neuro Imaging at the University of Southern California.

